# Undergraduate Pharmacology Written Question Papers of Different Universities of Bangladesh: Analysis of One Decade

**DOI:** 10.1101/2022.02.22.22271235

**Authors:** Fatema Johora, Asma Akter Abbasy, Sabiha Mahboob, Fatiha Tasmin Jeenia, Jannatul Ferdoush, Md Sayedur Rahman

**Affiliations:** Department of Pharmacology & Therapeutics, Army Medical College Bogura, Bogura; Department of Pharmacology& Therapeutics, Brahmanbaria Medical College, Brahmanbaria; Major Sabiha Mahboob, Medical Officer, CMH Bogura, Bogura; Department of Pharmacology& Therapeutics, Chattogram International Medical College, Chattogram; Department of Pharmacology & Therapeutics, BGC Trust Medical College, Chattogram; Department of Pharmacology, Bangabandhu Sheikh Mujib Medical University, Shahbag, Dhaka

**Keywords:** Undergraduate pharmacology, written question papers, SAQ, analysis

## Abstract

**Background:** It is expected that pharmacology education should prepare students as rational prescriber. Credibility of undergraduate pharmacology curricula is rather questionable in this aspect. As assessment shapes learning priorities, it is crucial to design assessment methods of pharmacology in right way to achieve the expected learning outcomes of future physicians.

**Materials and Methods:** This descriptive cross-sectional study was conducted to compare the reflection of curricular objectives, content coverage and national health priorities in undergraduate pharmacology written question papers (SAQ) of different universities of Bangladesh in last 10 years (January 2010 to November 2019). Total 131 question papers were collected, and reflection of curricular objectives, content coverage and selective disease burdens were evaluated and compared.

**Result:** One objective regarding factual knowledge (pharmacological effects, mechanisms of action, pharmacokinetic characteristics and adverse reactions of drugs) had significant higher weightage throughout the decade in all universities. There were statistically significant differences in weightage of reflection of five curricular objectives (p value <0.00001, 0.001, 0.003, 0.004, 0.02) among different universities. There was not a single question reflecting the ethical and legal issues involved in drug prescribing, development, manufacture and marketing in the decade in any university. Chemotherapy was the highest covered area (19.4±3.3), followed by central nervous system (16.0±3.4) and general principles of pharmacology (1 4.3±3.2) throughout the last ten years in all universities. There was statistically also significant difference among different universities in weightage of all content areas except Gastrointestinal pharmacology. Statistically significant difference (p value < 0.00001) among different universities in cardiovascular diseases burden was observed.

**Conclusion:** Current study found variation in pharmacology written question papers of different universities in the aspects of reflection of curricular objectives, content coverage and cardiovascular disease burden.

## INTRODUCTION

Undergraduate pharmacology education has always been a topic of tense debate. Credibility of pharmacology education in coping pace with rapid changes and requirements of clinical practice is questionable.^1, 2^ Studies concluded weakness in pharmacological knowledge and skill in prescribing medicines as an important factor of medication error.^3, 4, 5, 6^ Medical students around the world seek more requirement of attention in their pharmacology education.^7, 8^

Curriculum is a formal document including course content, objectives, learning outcomes, educational strategies and assessments for achievement of learning, teaching and evaluation under the guidance of educational institutions. It represents the expression of educational ideas in practices.^9, 10^ In any curriculum, a close match between instructional objectives, methods and assessment procedures are required.^11^ It is important to assure that objectives are measurable and specific level of competence being delineated. The assessment methods measures whether the objectives are achieved or not through formative and summative examinations.^12^ Formative assessments provide benchmarks to orient the learner who is approaching a relatively unstructured body of knowledge. They can reinforce students’ intrinsic motivation to learn and inspire them to set higher standards for themselves.^13^ On the other hand, summative assessments are intended to provide professional self-regulation and accountability as well as act as a barrier to further practice or training.^14^ All methods of assessment have strengths and intrinsic flaws.^15^ Written assessment has been considered as cornerstone of testing knowledge of graduates.^16^ Various types of questions like MCQ, SAQ, MEQ and SEQ can be used in written assessment.^17^ Due to limited validity, poor reliability and less objectivity, long essay questions lost popularity as instrument of written assessment.^18, 19^ Nowadays, SAQs are widely used because of greater objectivity, reliability, specificity and coverage of wider areas of course content.

In Bangladesh, first documented curriculum of MBBS program was introduced in 1988.^20^ After extensive review a revised curriculum was implemented on students of 2002-2003 session. Introduction of SAQ and MCQ in written assessment system instead of essay questions were the dramatic changes observed in that curriculum. ^21^ Later, reviewed MBBS curriculum 2012 has been developed, and implemented from session 2012-13.^22^ Seven public universities, Bangladesh University of Professionals (BUP), University of Dhaka (DU), University of Chittagong (CU), University of Rajshahi (RU), Shahjalal University of Science and Technology (SUST), University of Science and Technology, Chittagong (USTC) and Gono Bishwabidyalay (GB) are conducting MBBS examination of all medical students under the guidance of Bangladesh Medical & Dental Council (BMDC). After the implementation of curriculum 2002, Pharmacology had been assessed in 2nd professional exam since January, 2007 and in curriculum 2012, it has been assessed in 3rd professional exam since May, 2017. Several studies were done to evaluate Pharmacology curriculum, textbooks and question papers through different perspectives.^23-28^ In this background, current study was conducted to evaluate Pharmacology written question papers of different universities over last 10 years in the context of reflection of curricular objectives, content coverage and selective disease burdens.

## MATERIALS & METHODS

The objectives of this study were to compare the reflection of curricular objectives, content coverage and national health priorities in undergraduate pharmacology written question papers of different universities of Bangladesh.

### Study Design and Procedure

This descriptive cross-sectional study was conducted from January 2021 to June 2021. After obtaining ethical approval from the Institutional Review Board (IRB) of Combined Military Hospital (CMH) Bogura, researchers collected pharmacology written question papers (SAQ) of last 10 years (January 2010 to November 2019) of all 7 universities offering MBBS degree (Bangladesh University of Professionals, University of Dhaka, University of Chittagong, University of Rajshahi, Shahjalal University of Science and Technology, University of Science and Technology, Chittagong and Gono Bishwabidyalay). Total 131 question papers were collected and reviewed to meet the study objective.

Both curriculum 2002 and 2012 clearly described learning objectives for undergraduate pharmacology education. There were 10 curricular objectives in curriculum 2002 and 11 objectives in 2012.^21, 22^ Although there were some linguistic discrepancy in objectives of two curricula but essence of 10 objectives were same. Researchers sorted out curricular objectives suitable for assessing through written questions of professional examinations, and 7 objectives were selected. For convenience, researchers assigned chronological number against each objective, e.g. no. 1 objective, no. 2 objective, no. 3 objective etc. Then a list of probable questions for each curricular objective was prepared. For analysis, researchers reviewed the question paper thoroughly and identify questions reflecting specific objectives of curriculum. If present, then those questions were included and calculated for analysis. The Pharmacology written question papers (SAQ) contain total 84 marks with options, where students need to answer a maximum of 70 marks. Weightage was calculated as the number reflecting each objective out of 84 marks.

In undergraduate pharmacology curriculum, number of course contents were 11 (General principles of pharmacology, Autonomic pharmacology, Renal & cardiovascular pharmacology, Hematopietic pharmacology, Endocrine pharmacology, Gastrointestinal pharmacology, CNS pharmacology, Autacoids & drugs used in inflammation, Respiratory pharmacology, Chemotherapy and Clinical pharmacology). Course content wise allocation of wightage was also evaluated. Then, reflection of health care needs of community in the question papers was assessed by calculating how much weightage was given on disease burden of Bangladesh. According to Bangladesh Health Bulletin 2019, burden of cardiovacular disease was highest (30%), followed by communicable, maternal, perinatal and nutritional condition (26%), other non-communicable disease (12%), cancer (12%), Chronic respiratory disease (10%), injuries (7%) and diabetes (3%).^29^ A list of probable questions for pharmacotherapy of selective disease burdens (cardiovascular disease, chronic respiratory disease, cancer and diabetes) was prepared. Researchers reviewed the question papers thoroughly, and identified both direct and indirect questions related to pharmacotherapy of disease burdens, and weightage was calculated.

### Statistical analysis

Data was compiled, presented and and analyzed using Microsoft Excel 2007, and was expressed as mean percentage (standard deviation). One Way Analysis of Variance (ANOVA) was done to determine the significance of difference between the mean percentages. Statistical analysis was performed at a 95% confidence interval and significance was determined at p< 0.05.

## RESULTS

Total 131 SAQ papers (DU-20, CU-20, RU-20, SUST-19, USTC-16, GB-16 & BUP-20) of undergraduate pharmacology written question papers dated from January 2010 to November 2019 were analyzed. **Table I** showed that no. 1 objective had significant higher weightage throughout the last ten years in all universities. There were statistically significant differences in weightage of reflection of curricular objectives (No.1, 2, 4, 7 & 8) among different universities. There was not a single question reflecting no. 9 objective in last 10 years in any university.

**Table I:**
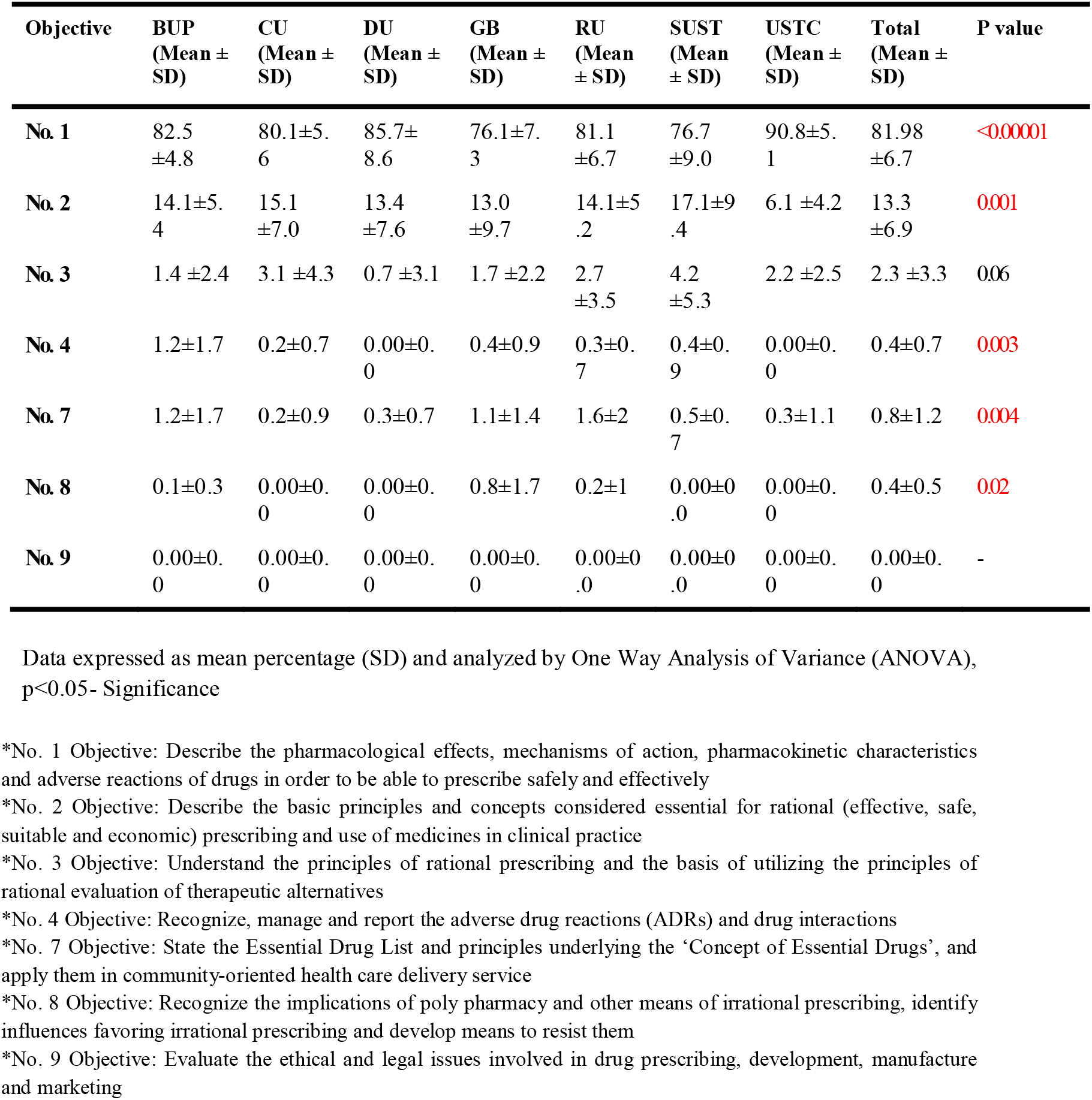
Objective-wise marks distribution.

**Table II** showed there were statistically significant differences in content-wise weightage among different universities except gastrointestinal pharmacology. Chemotherapy was the highest covered area, followed by central nervous system and general principles of pharmacology throughout the last ten years in all universities (Table II). In curriculum, allocated teaching hours for chemotherapy was highest, followed by general principles of pharmacology and central nervous system (Table III).

**Table II:**
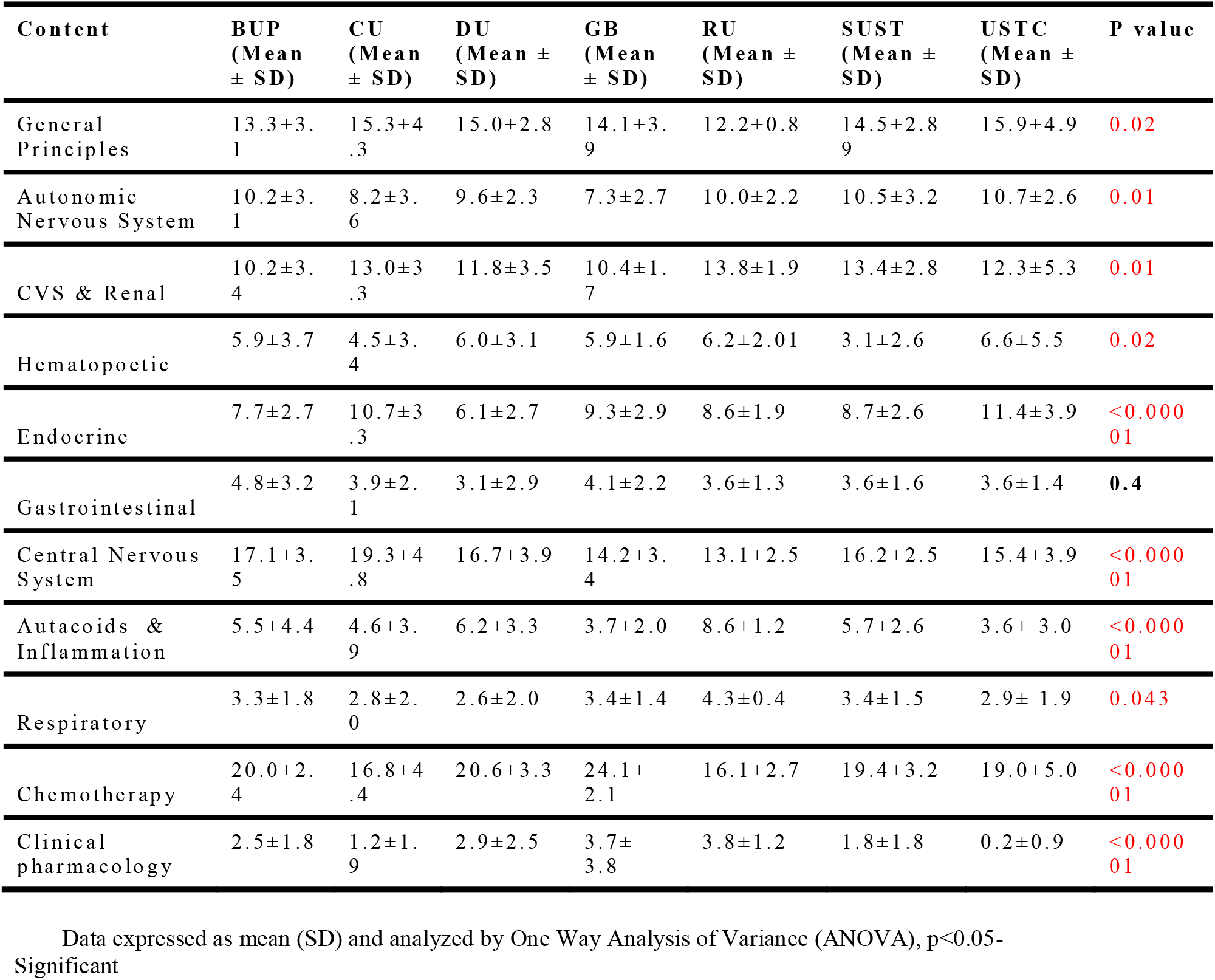
Content-wise marks distribution.

**Table III:**
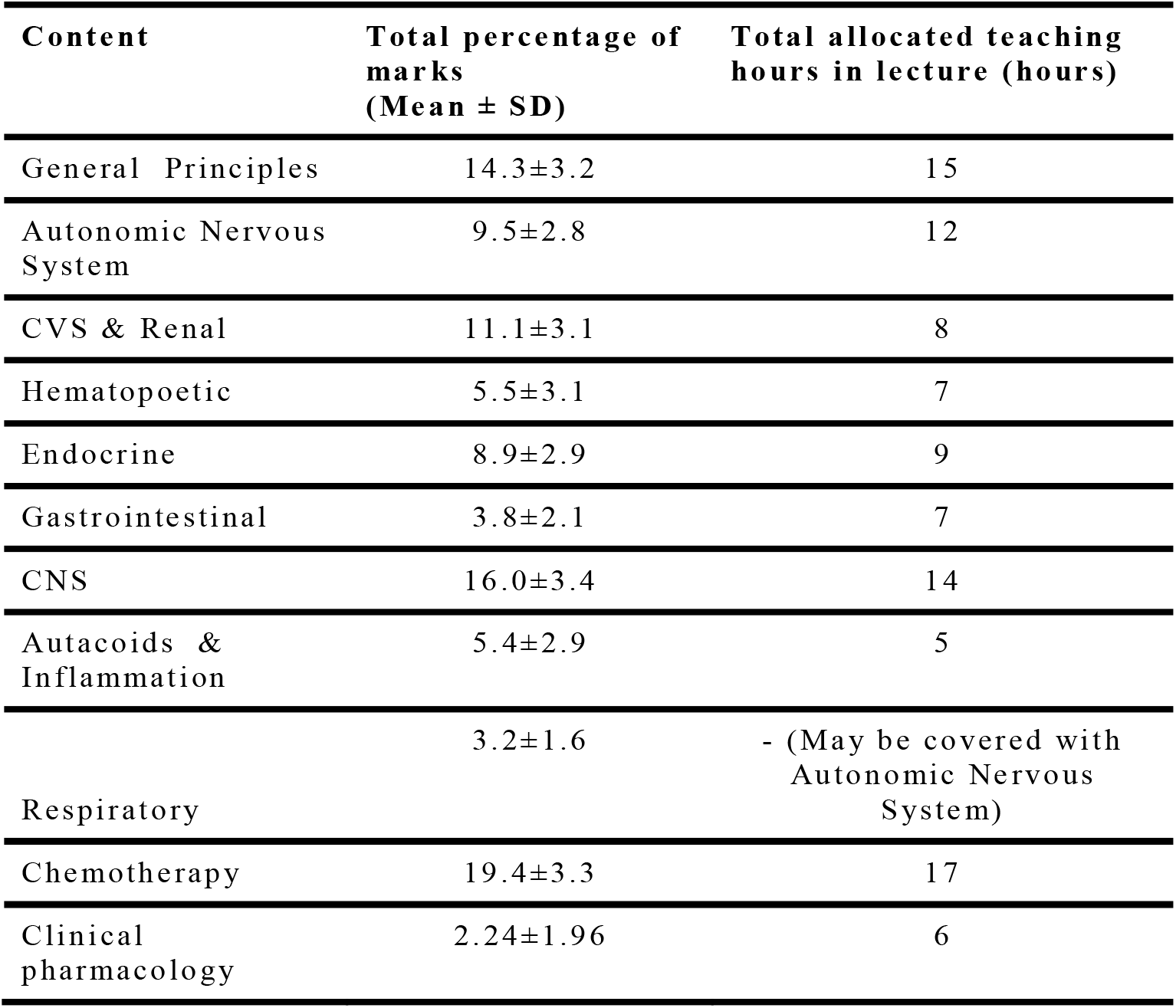
Content wise marks distribution and allocated teaching hours.

**Table IV** showed there was a statistically significant difference in weightage of reflection of cardiovascular disease burden among different universities.

**Table IV:** Disease burden wise marks distribution.

| Disease burden | BUP<br>(Mean ± SD) | CU<br>(Mean ± SD) | DU<br>(Mean ± SD) | GB<br>(Mean ± SD) | RU<br>(Mean ± SD) | SUST<br>(Mean ± SD) | USTC<br>(Mean ± SD) | Total<br>(Mean ± SD) | P value |
| --- | --- | --- | --- | --- | --- | --- | --- | --- | --- |
| Cardiovascular (30%) | 15.8±3.2 | 16.1±3.9 | 19.6±5.0 | 14.3±3.8 | 20.5±3.8 | 18.4±3.9 | 19.5±5.3 | 17.7±4.1 | **<0.00001** |
| Cancer (12%) | 0.2±1.0 | 0.0±0.0 | 0.4±1.1 | 0.0±0.0 | 0.5±1.2 | 0.0±0.0 | 0.0±0.0 | 0.5±0.5 | 0.3 |
| Chronic respiratory disease (10%) | 3.2±1.9 | 3.1±2.0 | 2.7±2.4 | 3.8±2.2 | 4.3±0.4 | 4.1±1.9 | 3.5±1.8 | 3.5±1.8 | 0.2 |
| Diabetes (3%) | 3.8±1.9 | 4.0±1.2 | 3.3±3.2 | 4.5±1.6 | 4.0±1.5 | 3.8±2.0 | 5.3±2.7 | 4.1±2.0 | 0.5 |

Data expressed as mean (SD) and analyzed by One Way Analysis of Variance (ANOVA), p<0.05-Significant

## DISCUSSION

Re-addressing assessment methods is a crucial step for transformation of medical education and over the years, innovative measures are taken for provision of accurate and timely assessment of future physicians.^30^ Current study was conducted to analyze and compare pharmacology question papers of different universities of Bangladesh in respect to reflection of curricular objectives, content coverage and selective disease burdens for last 10 years.

Objectives are statements that describe the end-points or desired outcome of the curriculum which guide the assessment method, outline content and instructional materials, contribute towards achieving the intended educational outcomes, shaping expectations, prepare learners for the educational activity and the standard by which their performance will be measured.^31^ No. 1 objective had significant higher weightage throughout the last ten years in all universities, followed by no. 2 and no. 3 objective. No. 1 objective actually presents knowledge and comprehension, whether no.2 and no. 3 objectives demonstrate application, analysis and evaluation. Higher proportion of coverage of factual knowledge in professional exam question papers of pharmacology was also revealed in studies conducted in Bangladesh and India.^24, 26, 28, 32, 33^ Important issues of prescribing like adverse drug reactions, essential medicines, rational prescribing, ethical and legal aspects are still neglected in undergraduate pharmacology education, and this finding is concordance with previous literatures.^23,27^ It is assumed that it’s convenient for question setters and moderators to prepare questions in traditional way as they are probably not adequately trained in adaptation of changing aspects of pharmacology teaching-learning.^34^ Current study also found statistically significant difference of weightage in reflection of curricular objectives (No.1, 2, 4, 7 & 8) among different universities, and there was no available research to compare the findings.

In case of content coverage, it was found that Chemotherapy, CNS pharmacology, General principles of pharmacology, Renal & cardiovascular pharmacology and Autonomic pharmacology were the maximum weightage of content area, and the findings were similar to related studies conducted in Bangladesh.^26, 28^ Increased weightage on Hematopietic pharmacology was observed in current study that was similar to Begum et al^24^ but contrary to two recent studies.^26, 28^ Weightage on Endocrine pharmacology was consistent with related studies.^23, 24, 26, 28^ There was statistically significant difference among different universities in weightage of all content areas except Gastrointestinal pharmacology, and no comparable study was found to correlate this finding. There were some discrepancies in weightage of different contents with their allocated teaching hours, and that was concordance with an earlier study conducted in Bangladesh.^23^ A test blueprint and question bank would be helpful for the question setters and moderators for adequate content coverage.^28, 35^

Current study compared reflection of selective disease burdens in written question papers and found statistically significant difference (p value <0.00001) in cardiovascular diseases among different universities. Although there was no comparable study, earlier researches concluded less emphasis on national health priorities in pharmacology question papers of Bangladesh.^23, 24^ One recent study conducted in Bangladesh revealed pharmacology written question papers of different universities under guidance of same curriculum of BMDC differ in the aspect of weightage of problem-based questions.^36^

## CONCLUSION

Current study revealed variation in undergraduate pharmacology written question papers of different universities in the aspects of reflection of curricular objectives, content coverage and cardiovascular disease burden. A clear direction of weightage of each objective, contents and disease burdens are needed to mention in the undergraduate curriculum. As assessment drives learning priority, it’s crucial for question setters and moderators to maintain uniformity of written question papers of different universities following same curriculum of BMDC.

## Data Availability

All data produced in the present study are available upon reasonable request to the authors

## Notes

### Competing Interest Statement

The authors have declared no competing interest.

### Funding Statement

This study did not receive any funding

### Author Declarations

Ethical approval was obtained from the Institutional Review Board (IRB) of Combined Military Hospital (CMH) Bogura

